# Estimating the prevalence of undiagnosed Long COVID using health insurance claims data

**DOI:** 10.64898/2026.11.24.26368867

**Authors:** Elise N. Grover, Andrew Hill, Emma J. Gunn, Rick DeVoss, Rifei Liang, Sarah E. Jolley, Marcelo C. Perraillon, Elizabeth J. Carlton

## Abstract

**Background:** Long COVID (LC) is a complex, multisystemic condition whose prevalence is poorly understood due, in part, to underdiagnosis.

**Methods:** We used the Colorado All Payer Claims Database (CO APCD), which covers approximately 70% of insured Coloradans, to train a machine learning eXtreme Gradient Boosting model to predict undiagnosed LC. The 243 candidate predictors included clinical symptoms, comorbid conditions, healthcare utilization, pharmaceutical prescriptions, and individual- and neighborhood-level socioeconomic conditions. Models were trained on a dataset of 38,064 adults (18+), including individuals with the LC diagnostic code (U09.9), and a set of comparators without the code, matched by age, sex and payer. The trained model was used to estimate the prevalence of undiagnosed LC within a larger CO APCD extract comprising 2.1 million adults. We compared diagnostic patterns across sociodemographic and geographic characteristics by calculating the ratio of predicted undiagnosed to diagnosed prevalence.

**Results:** The prevalence of undiagnosed adult LC in Colorado was estimated to be 8%, corresponding with an estimated 7 undiagnosed cases for every 1 diagnosed case. High levels of healthcare utilization and respiratory-related symptoms were top predictors of LC. Conditions that helped to rule out LC (e.g., musculoskeletal pain) were also important to model performance. Groups with higher proportions of probable, undiagnosed LC relative to diagnosed LC include younger adults, males, and urban populations.

**Conclusions:** Leveraging medical claims, neighborhood socioeconomic and geographic data in advanced machine learning algorithms can facilitate rapid estimation of the hidden burden of undiagnosed LC and identify populations less likely to receive a diagnosis.

**Significance Statement:** Long COVID is a multisystemic, disabling condition that is underdiagnosed, making it difficult to align healthcare resources with patient needs. We developed an optimized machine learning algorithm using medical claims data to identify high probability, undiagnosed Long COVID cases. We used this algorithm to estimate the prevalence of undiagnosed Long COVID and evaluate differences in diagnosis by clinical features and sociodemographic factors. These approaches demonstrate how healthcare utilization patterns can be used to identify key predictors of Long COVID and monitor the distribution of complex and emerging diseases that are not well described by existing surveillance systems.

## Introduction

Post-acute sequelae of coronavirus disease 2019 (COVID-19), commonly known as “Long COVID” (LC), is a complex, multisystemic condition characterized by symptoms that persist after infection with severe acute respiratory syndrome coronavirus 2 (SARS-CoV-2). Although the first reports of long-term symptoms appeared within months of SARS-CoV-2’s emergence (1), a diagnostic code (U09.9) wasn’t introduced until October 2021 (2), and a consensus definition was not established until 2024, when the National Academies of Sciences, Engineering, and Medicine (NASEM) defined LC as an infection-associated chronic condition that “occurs after SARS-CoV-2 infection and is present for at least 3 months as a continuous, relapsing and remitting, or progressive disease state that affects one or more organ systems” (3). LC is recognized as a condition that can impair quality of life, reduce the ability to participate in the workplace, and incur substantial costs to the healthcare system. A key priority is ensuring that there are resources to meet the needs of LC patients, which necessitates timely and accurate diagnoses and estimates of population-level LC prevalence.

Despite an established diagnostic code and consensus definition, a substantial proportion of LC goes undiagnosed. Studies highlight that medical skepticism, knowledge gaps and inexperience have led to misdiagnoses across a range of healthcare settings (4–7). LC diagnostic code usage also varies substantially by region and healthcare facility, and typically only captures the most severe cases (8–10). The lack of objective biomarkers or diagnostic tests compounds these challenges, requiring providers to rely on self-reported symptoms and a time-consuming process of elimination before diagnosis is considered (4, 11, 12). This process is further complicated by inconsistent and sometimes episodic symptoms that fluctuate in presentation and severity over periods of days to months (12–14). Additionally, LC symptoms frequently overlap with other nonspecific conditions (e.g., fibromyalgia, chronic fatigue syndrome) (5, 15), and can exacerbate underlying conditions, all of which can lead to diagnostic delays and missed diagnoses (14, 16). Missed or delayed diagnosis can impair patient access to care, and at a population level, makes it challenging to estimate the prevalence of LC in populations.

Estimates of LC prevalence range widely, from 6% to over 15%, with higher estimates generally derived from self-reported data (17–21). Our prior work and studies by others have highlighted the gap between LC diagnosis in medical records and probable LC based on symptom surveys and self-reports. Studies in the United Kingdom have estimated that approximately one in every 15 LC cases received a diagnosis in the early years of the COVID-19 pandemic (17, 19). Similarly, whereas our team previously found that less than 1% of Colorado adults had an LC diagnostic code in their medical record between October 2021 and August 2022 (22), a nationally representative survey that used self-report put the Colorado LC prevalence between 5.8 and 7.0% in 2022 (21).

Additionally, there is suggestive evidence that underdiagnosis of LC is not uniformly distributed across the population. Instead, LC disproportionately affects individuals who experience greater sociodemographic disadvantage (20), which, in turn, can lead to a disproportionately lower likelihood of diagnosis (23).

We set out to estimate the undiagnosed prevalence of LC across Colorado to inform state efforts to improve access to LC care, and to assess whether some groups were more or less likely to receive a diagnosis. We used a predictive framework that combined medical claims data with machine learning (ML) approaches appropriate for high-dimensional data. Pairing ML models with medical record databases has proven to be a fruitful method of creating high-performing prediction models that have clarified the symptom profiles of known LC cases (24–30). We are aware of two prior attempts to use such algorithms to estimate LC prevalence among people with prior SARS-CoV-2 infection in the United States (28) and Canada, the latter of which also estimated the undiagnosed gap to be >10x the number of diagnosed cases (26). We build on that work by 1) estimating LC in the full population, removing the requirement of prior acute COVID-19 given the shift toward at-home testing and more mild infections that may not prompt care-seeking behavior; 2) evaluating the ratio of undiagnosed to diagnosed LC across socio-demographic groups.

In this study, we developed a predictive ML algorithm using the Center for Improving Value in Health Care (CIVHC) Colorado All Payer Claims Database (CO APCD), which offers high-dimensional, longitudinal data for over 70% of the Colorado population with medical coverage, representing approximately 3.8 million individuals (31). As the repository of health insurance claims information for Colorado residents covered by Medicare, Medicaid and 33 commercial payers, the CO APCD is the most comprehensive source of medical, pharmacy, and dental claims in the state (32). We compiled claims-based documentation of symptoms and comorbidities, healthcare utilization, prescription fills, and individual demographics from the CO APCD for the years 2021 – 2023, and used claims-based location to estimate neighborhood socioeconomic conditions. Using extracts of our compiled data, we trained, validated and applied a predictive algorithm for identifying probable LC and estimated the prevalence of undiagnosed LC in Colorado for the years 2022 and 2023, regardless of SARS-CoV-2 infection history. We also used the CO APCD to estimate the total prevalence of LC and compared characteristics of diagnosed and undiagnosed populations to better understand whether subgroups of the population are at higher risk of being undiagnosed. Together, our findings shed light on the hidden burdens and non-uniform distribution of LC diagnoses, information that can be used to improve access to care for LC patients. Our methodological framework has potential extensions for estimating the distribution of other complex diseases and syndromic conditions.

## Results

### Characteristics of the training sample

The training dataset included 12,688 adults (age 18+) with an LC diagnosis (U09.9) and 25,376 comparators without, matched on age, sex, and payer. A comparison of selected characteristics for those with and without an LC diagnosis is provided in Supplementary Table S1. In brief, the LC diagnosed population had more than twice the proportion of respiratory-related claims (e.g., respiratory signs and symptoms: 83.3% vs 40.0%; respiratory failure: 29.3% vs 7.2%; pneumonia: 31.6% vs 5.8%; asthma: 27.2% vs 11.7%; COPD: 18.1% vs. 8.8%), as well as more comorbidities and greater healthcare utilization than the comparators. For example, chronic pulmonary disease was more than twice as common in the LC diagnosed population (41.7% vs 19.8%), and having at least one hospital visit was almost three times as likely in the LC diagnosed population (22.3% vs 8.4%). The LC diagnosed population also had more prescription medication claims, including corticosteroids (53.8% vs. 20.7%) and bronchodilators (54.3% vs. 18.5%) than the undiagnosed comparators. Individuals with an LC diagnosis were less likely to reside in rural areas (13.1% vs 10.8%), and more likely to have a lower median income ($76,394 vs $79,147) than their matched comparators.

### Training model performance and feature importance

Our primary model, the all-predictors model, included 243 predictors representing clinical symptoms, comorbid conditions, healthcare utilization, pharmaceutical prescriptions, individual and neighborhood sociodemographics (Supplementary Table S2). To select a final probability threshold to use for classifying predictions output by our ML algorithm, we compared model performance metrics (the median and 95% confidence intervals (CIs) of precision, recall, and F1 score across 1,000 bootstrap samples of the training data) across a set of potential classification thresholds ranging from ≥50% to ≥90% (Supplementary Table S3). We selected a final threshold of ≥80% to categorize individuals as probable LC, as this yielded excellent precision (≥90%) while maintaining an acceptable recall (≥50%). Bootstrapped iterations of feature importance rankings from the all-predictors model highlighted that the number of office visits and respiratory signs and symptoms were consistently the best predictors of LC, ranked first and second respectively (Supplementary Figure S1).

### Performance of the final model and sensitivity models

The final (non-bootstrapped) all-predictors model had a precision of 90.3, recall of 56.1, F1 score of 69.2 and area under the precision-recall curve (AUC-PR) of 87.4 (Table 1). The “Expert-Informed” model, in which clinical symptom predictors were limited to those identified by literature review and clinical experts (”Expert-Informed” Model), had modestly lower performance: precision was modestly higher (91.9) but all other performance indicators were lower than those of the all-predictors model. Our sensitivity analysis to assess whether the all-predictors model was biased toward the sickest patients indicated that when healthcare utilization, prescription medications, and symptoms were included as binary rather than count variables (“Binary” Model), the model had high precision (91.2), but the lowest recall, F1 score, and AUC-PR of any model. Similarly, models including a parsimonious set of predictors by restricting to the top 50 (”Top 50” Model) and top 25 predictors (”Top 25” Model) had similarly high precision, but the recall, F1 score and AUC-PR were lower than those of the all-predictors model. In a final “Temporal” model designed to evaluate the stability of our all-predictors model over time, performance was highest when later data was used for training: the model trained on the latest available data (Training on Folds 2-4, testing on Fold 1) performed the best (Precision: 90.4, Recall: 65.6), while the model trained on the earliest data (Training on Folds 1-3, testing on Fold 4) performed the worst (Precision: 88.8, Recall: 61.0).

**Table 1.** Training model performance comparison between the all-predictors model and the sensitivity analysis models, using an 80% probability threshold.

| **Model** | **Precision** | **Recall** | **F1 Score** | **AUC-PR** |
|---|---|---|---|---|
| All-predictors model<sup>†</sup> | 90.3 | 56.1 | 69.2 | 87.4 |
| Sensitivity model A: Expert-informed model | 91.9 | 48.0 | 63.1 | 86.3 |
| Sensitivity model B: Binary model | 91.2 | 32.0 | 47.3 | 80.0 |
| Sensitivity model C: Top 50 predictors | 90.6 | 48.8 | 63.4 | 85.6 |
| Sensitivity model D: Top 25 predictors | 90.0 | 43.5 | 58.6 | 84.2 |
| Sensitivity model E: Temporal model (all folds)<sup>‡</sup> | 89.4 | 57.6 | 70.1 | 86.8 |
| *Sensitivity model E: Test on Temporal Fold 1* | *90.4* | *65.6* | *76.0* | *89.9* |
| *Sensitivity model E: Test on Temporal Fold 2* | *88.9* | *58.4* | *70.5* | *86.8* |
| *Sensitivity model E: Test on Temporal Fold 3* | *89.1* | *57.5* | *69.9* | *86.3* |
| *Sensitivity model E: Test on Temporal Fold 4* | *88.8* | *46.4* | *61.0* | *83.1* |

<sup>†</sup> To make the output performance metrics of the "all-predictors" model directly comparable to the sensitivity models included here, bootstrapped iterations training the model on repeat random samples of the training data were not included. Instead, a single training iteration was performed using the entire 75% training dataset, and model performance metrics were calculated using the reserved 25% testing data. This difference accounts for the slightly higher model performance reported here, as compared to the bootstrapped model performance reported in Table 2 wherein training was conducted on random bootstrapped samples of the training dataset.
<sup>‡</sup> To get a summary estimate of model performance across all four folds, true positives, true negatives, false positives and false negatives were summed across all four cross-validation iterations (each using three folds for training, and one-fold for testing) to create a summary of performance across all folds. The performance of the model when testing on each successive fold (and training on the three other available temporal folds) is also provided in Table 3 above to illustrate changes in model performance over time.

**Table 2.**
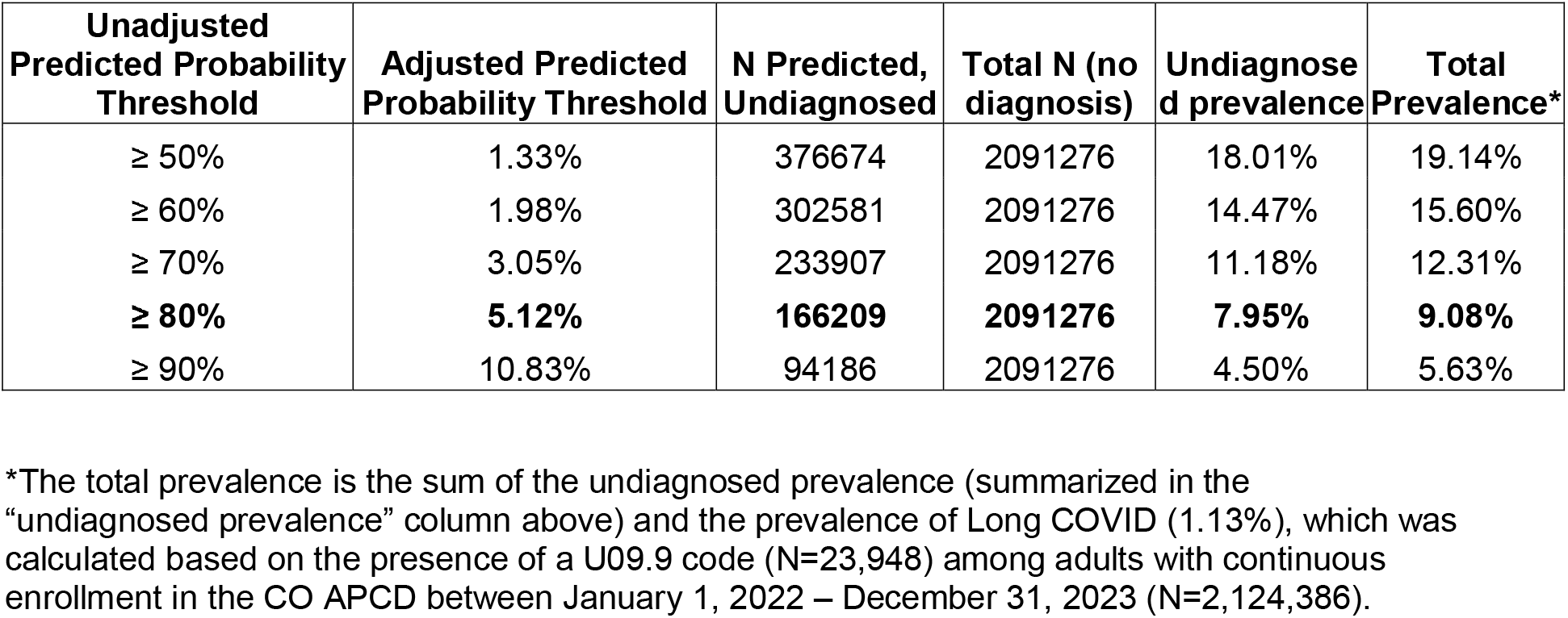
Estimated prevalence of diagnosed and undiagnosed long COVID among the 2,129,355 people in the Colorado All Payer Claims Database (APCD) in Colorado between 01/01/2022 and 12/31/2023 (training and testing datasets) using our predictive algorithm.

| **Unadjusted Predicted Probability Threshold** | **Adjusted Predicted Probability Threshold** | **N Predicted, Undiagnosed** | **Total N (no diagnosis)** | **Undiagnosed prevalence** | **Total Prevalence\*** |
|---|---|---|---|---|---|
| ≥ 50% | 1.33% | 376674 | 2091276 | 18.01% | 19.14% |
| ≥ 60% | 1.98% | 302581 | 2091276 | 14.47% | 15.60% |
| ≥ 70% | 3.05% | 233907 | 2091276 | 11.18% | 12.31% |
| **≥ 80%** | **5.12%** | **166209** | **2091276** | **7.95%** | **9.08%** |
| ≥ 90% | 10.83% | 94186 | 2091276 | 4.50% | 5.63% |

\*The total prevalence is the sum of the undiagnosed prevalence (summarized in the “undiagnosed prevalence” column above) and the prevalence of Long COVID (1.13%), which was calculated based on the presence of a U09.9 code (N=23,948) among adults with continuous enrollment in the CO APCD between January 1, 2022 – December 31, 2023 (N=2,124,386).

### Predictor importance in the final validated model

In the final all-predictors model, healthcare utilization patterns and respiratory symptoms represented a larger proportion of the top 15 predictors than what would be expected by chance alone (Figure 1). Healthcare utilization patterns represented 13.3% of the top 15 predictors (i.e., number of office visits, ranked 1^st^; number of ER visits, ranked 6^th^), despite comprising only 1.6% of the predictors included in the model. Similarly, whereas respiratory symptoms made up 4.1% of the predictors in the model, they comprised 20% of the top 15 predictors, including respiratory signs and symptoms (ranked 2^nd^), non-tuberculosis caused pneumonia (ranked 7^th^) and respiratory failure, insufficiency or arrest (ranked 13^th^). Pharmaceutical claims were also informative, with antiasthma bronchodilators and adrenal corticosteroids ranked 5^th^ and 11^th^, respectively. In most cases, the top-performing predictors were positively associated with LC status, indicated in Figure 1 using red to the right of the central zero vertical line and blue to the left. However, 4 of the top 15 predictors represented clinical symptom codes that were more informative for ruling out LC: exposure, screening or contact with infectious disease (ranked 6th), neoplasm-related encounters (ranked 7th), medical examination or evaluation (ranked 9th), and musculoskeletal pain (ranked 15th).

**Figure 1.**
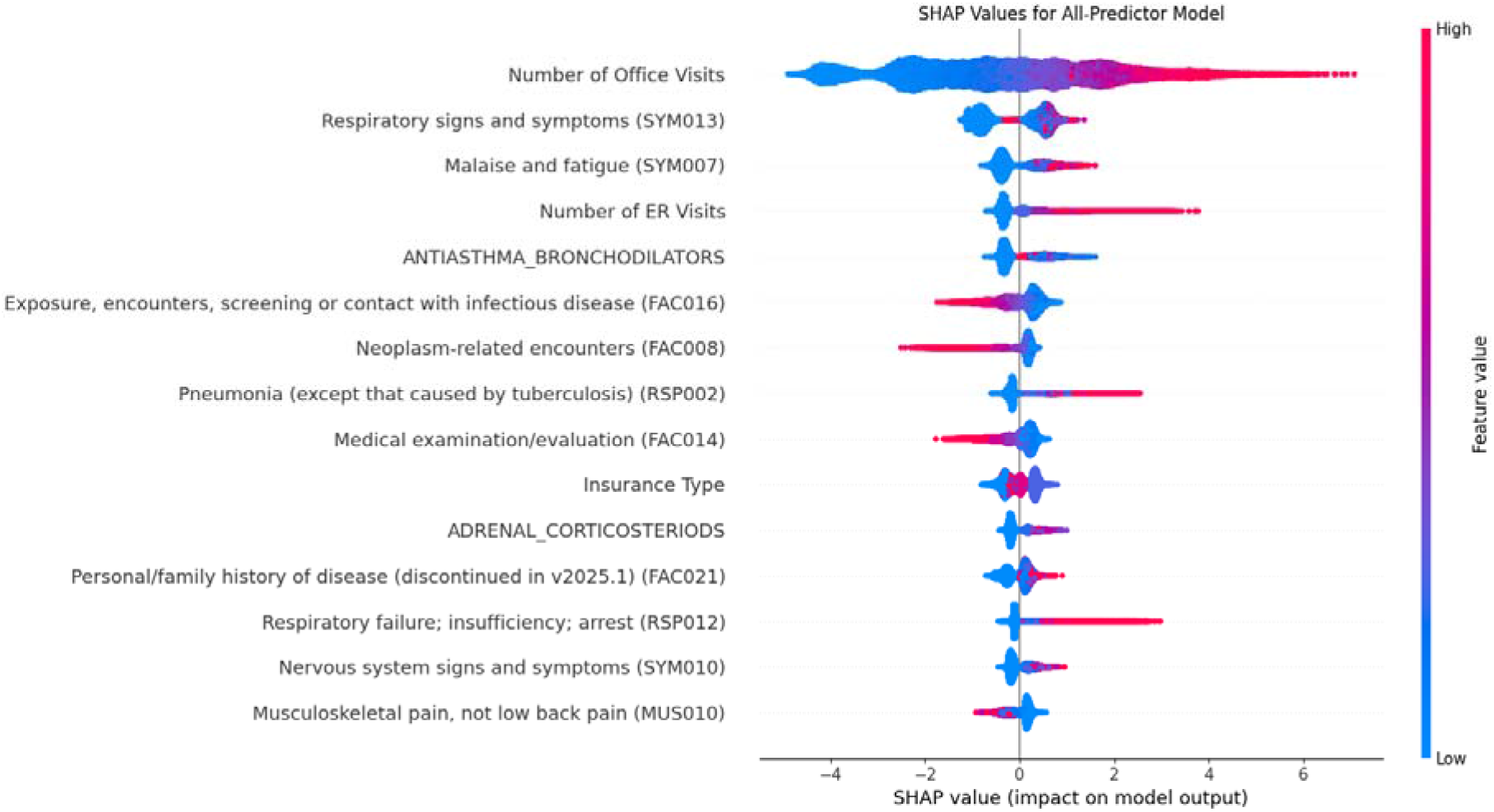
SHapley Additive exPlanations (SHAP) Beeswarm plot using the training data in the final all-predictors model (i.e., the model that included all 243 candidate predictors).The SHAP value indicates the degree of influence that a predictor has on pushing the model’s predictions up or down, relative to the global average. The y-axis of the plot depicts the final rank of the top 15 performing predictors from the model ranked according to their mean absolute SHAP value, with the top predictor representing the best performing predictor (number of office visits), and the bottom the 15^th^ best performing predictor (musculoskeletal pain). The distribution of influence that each of the 15 top performing predictors had on the prediction of Long COVID diagnosis is shown on the x-axis. Red indicates higher feature values, while blue indicates lower feature values. Negative SHAP values (to the left of the zero line, representing the baseline average) indicate a decrease in the probability of a long COVID diagnosis, while positive SHAP values (to the right of the baseline average zero line) indicate an increase in the probability of a long COVID diagnosis.

### Variable rank stability comparison between final model and sensitivity models

A comparison of variable ranks between the all-predictors model and the five sensitivity models demonstrated similar variable ranks for all but the binary model, which showed more variability (Supplementary Table S4). For example, whereas all other models ranked the number of office visits 1^st^, and essential hypertension claim counts among their top 20 predictors, the binary model ranked office visits (any claims vs. none) as 7^th^ and essential hypertension (any claims vs. none) 158^th^. Nevertheless, nearly all other variables ranked among the top 10 in other models ranked in the top 20 for the binary model.

To investigate whether variable influence changed over time, we compared variable ranks across the four temporal folds of Sensitivity Model E (Supplementary Table S5). Whereas we found minimal variability among the ranks of the top 15 predictors, for predictors ranked >15, Fold 1 tended to be the most distinct, relative to Folds 2-4.

### Estimating LC prevalence

The observed prevalence of the LC diagnostic code in the target population was 1.13%, which comprised 23,948 adults (18+) with an LC diagnosis code (U09.9) who were continuously enrolled in the CO APCD between 2022-2023 (N=2,124,386).

We estimate that the burden of undiagnosed LC in Colorado is 8.0% (Table 3), or approximately 7 times the observed prevalence of clinically diagnosed LC within our cohort of continuously enrolled adults in the CO APCD between 2022-2023 (1.13%), and almost 12 times the prevalence of diagnosed LC estimated for the broader Colorado population (0.67%) (22). Our estimate is based on our high-probability threshold (80%). Prevalence estimates ranged from 4.5 to 18% using probability thresholds from ≥50 to ≥90%.

**Table 3.** Comparison of populations with diagnosed Long COVID and those predicted to have Long COVID who lack a diagnosis by comorbid conditions, symptoms, healthcare utilization and pharmaceutical prescriptions.

| **Population Characteristics** | **Diagnosed Sample** |  | **Predicted, Undiagnosed\*** |  |
|---|---|---|---|---|
|  | **23,948** |  | **166,209** |  |
| **Symptoms** | **N** | **%** | **N** | **%** |
| Malaise & fatigue | 7,217 | 55.0% | 80,438 | 48.4% |
| Sleep wake disorders | 6,529 | 49.7% | 74,555 | 44.9% |
| Respiratory symptoms | 10,231 | 77.9% | 135,940 | 81.8% |
| Nervous system symptoms | 6,597 | 50.2% | 77,273 | 46.5% |
| Exposure to infectious disease | 10,488 | 79.9% | 125,231 | 75.3% |
| Esophageal disorders | 5,163 | 39.3% | 59,058 | 35.5% |
| Nonspecific chest pain | 4,575 | 34.8% | 56,848 | 34.2% |
| Respiratory failure; insufficiency | 2,813 | 21.4% | 32,563 | 19.6% |
| Fluid and electrolyte disorders | 3,648 | 27.8% | 46,433 | 27.9% |
| Pneumonia | 2,477 | 18.9% | 33,080 | 19.9% |
| Immunity disorders | 984 | 7.5% | 8,591 | 5.2% |
| Asthma | 3,441 | 26.2% | 43,294 | 26.1% |
| COPD | 2,712 | 20.7% | 30,422 | 18.3% |
| Musculoskeletal pain (not low back) | 8,978 | 68.4% | 104,604 | 62.9% |
| Other specified connective tissue disease | 4,178 | 31.8% | 43,010 | 25.9% |
| **Comorbidities** | **N** | **%** | **N** | **%** |
| Myocardial infarction | 1,572 | 6.6% | 10,801 | 6.5% |
| Chronic pulmonary disease | 10,216 | 42.7% | 71,098 | 42.8% |
| Congestive heart failure | 3,202 | 13.4% | 23,782 | 14.3% |
| Renal Disease | 3,872 | 16.2% | 27,496 | 16.5% |
| Connective tissue disease | 1,548 | 6.5% | 9,980 | 6.0% |
| **Healthcare Utilization** | **N** | **%** | **N** | **%** |
| ≥1 Hospitalization | 3,096 | 23.6% | 40,588 | 24.4% |
| ≥1 ER visit | 7,804 | 59.4% | 115,377 | 69.4% |
| ≥1 specialist visit | 10,890 | 82.9% | 138,534 | 83.3% |
| ≥1 Office visit | 12,878 | 98.1% | 165,380 | 99.5% |
| **Prescriptions** | **N** | **%** | **N** | **%** |
| ≥1 Adrenal corticosteroids | 5,285 | 40.3% | 81,403 | 49.0% |
| ≥1 Antiasthma bronchodilators | 5,264 | 40.1% | 80,796 | 48.6% |
| ≥1 Prescriptions | 10,600 | 80.7% | 134,620 | 81.0% |

### Comparing characteristics of predicted, undiagnosed LC to diagnosed LC

The predicted, undiagnosed population was similar to the diagnosed LC population when comparing claims-based symptoms, comorbidities and healthcare utilization (Table 3). One exception to this: the proportion of patients with at least one ER visit was higher among the predicted undiagnosed group (69.4%) as compared to the diagnosed LC group (59.4%). The predicted undiagnosed group also had a higher prevalence of adrenal corticosteroids (49.0% vs. 40.3%) and antiasthma bronchodilators (48.6% vs. 40.1%) prescriptions, relative to the diagnosed LC group.

The ratio of predicted, undiagnosed to diagnosed LC varied by age, sex and payer, noting that a higher ratio indicates a greater estimated relative burden of undiagnosed LC (Figure 2, Supplementary Table S6). The ratio was higher for men than women, was highest for the youngest adults (12.45, 95% CI 11.95, 12.95), and was also high for the oldest adults (80+ 7.69, 95% CI 7.39, 8.01). People with dual insurance (Medicare and Medicaid) had the lowest ratio (2.76, 95% CI 2.62, 2.90), suggesting higher detection rates in this population. American Indians/Alaskan Natives had the lowest ratio (5.62, 95% CI 4.77, 6.69) of all race categories. There were no other major differences in diagnostic patterns by ethnicity or race, however, given the high rates of missingness of race in the CO APCD, these findings should be interpreted with caution. The undiagnosed to diagnosed ratio also increased with population density, suggesting a greater proportion of undiagnosed LC among urban populations, relative to those residing in rural or frontier regions.

**Figure 2.**
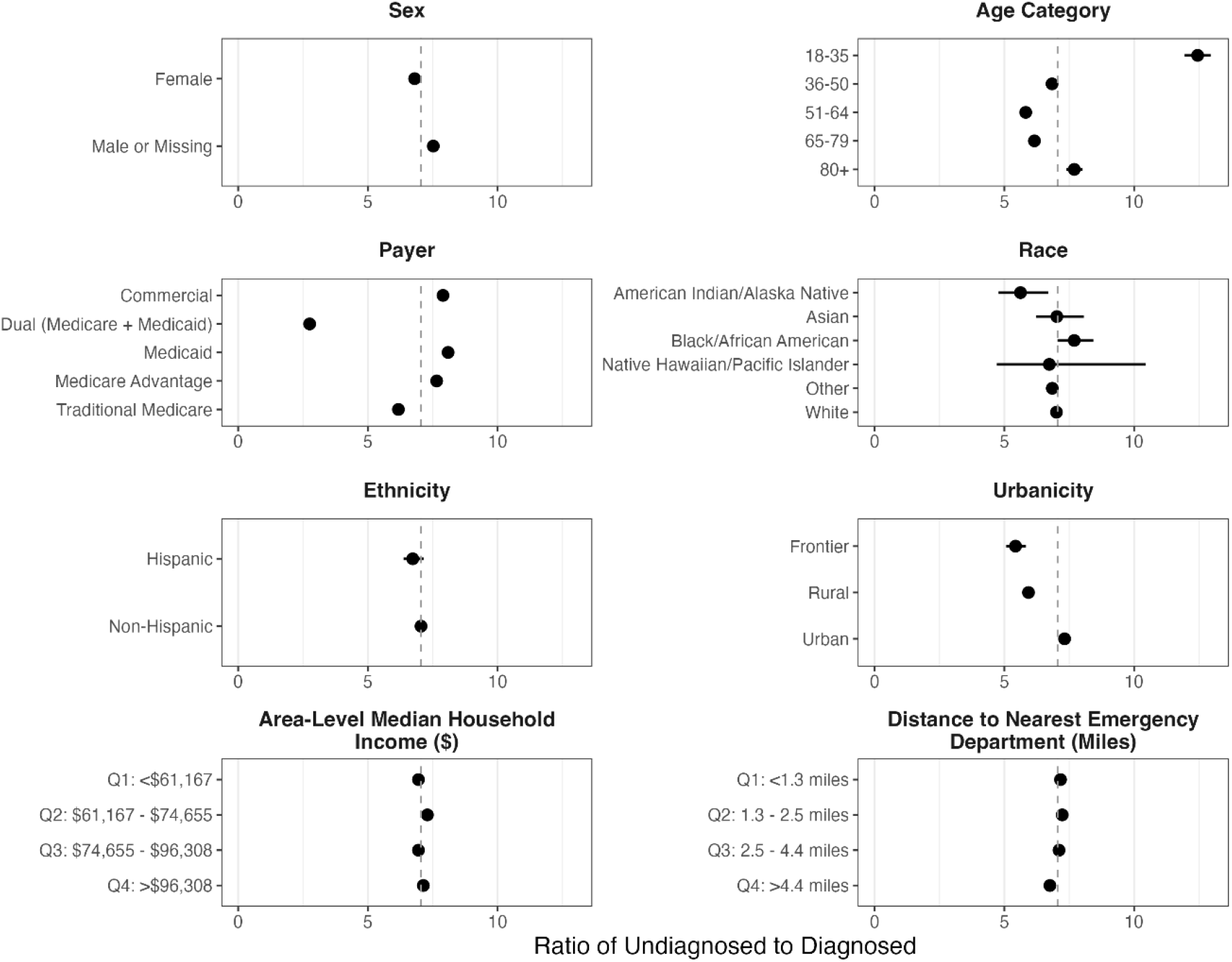
Participant sociodemographic characteristic categories (y-axis) by the ratio of predicted, undiagnosed Long COVID (LC) to diagnosed LC (x-axis) for sex, age category, payer, race, ethnicity, urbanicity, median household income and distance to the nearest emergency department. Dots indicate the point estimate of the ratio, while horizontal lines indicate the 95% confidence interval, based on 1,000 bootstrap iterations. The gray dotted line at 7.05 on the x-axis represents the undiagnosed-to-diagnosed prevalence ratio across the entire population. Dots that are far to the left or right of this dotted line represent deviations from the group norm, highlighting potential inequities.

## Discussion

In this analysis, we demonstrate that an optimized machine learning algorithm can be a timely tool for leveraging medical claims data to advance the state of LC surveillance. We expand on the scientific body of evidence documenting the set of characteristics typical of diagnosed LC cases, and harness this information to identify high probability, undiagnosed LC cases in the population, which we estimate to occur at a frequency of approximately 7 undiagnosed cases for every one diagnosis case in Colorado. Among the top predictors of an LC diagnosis were healthcare utilization intensity indicators, as well as several respiratory-related symptom codes. We demonstrate how this algorithm can be applied to estimate undiagnosed LC prevalence within and across sociodemographic groups and introduce ratio-based comparisons of probable, undiagnosed LC to diagnosed LC, highlighting the populations most or least likely to receive an LC diagnosis when they have a combination of symptoms, healthcare utilization patterns, comorbidities and pharmaceutical prescriptions consistent with LC. Unlike most studies aiming to estimate the burden of LC, we did not rely on documentation or reports of prior SARS-CoV-2 infection, freeing us from an increasingly common source of error given increasing home-testing usage and mild or undetected initial infections.

A central challenge of estimating LC prevalence is the absence of a gold standard diagnostic tool, which may be a key source of underdiagnosis. Our work required assumptions about the alignment between the LC diagnostic code and true LC cases and non-LC individuals that are difficult to verify. Below, we evaluate the evidence that our model is producing results that show consistency and coherence, discuss the implications of our assumptions and areas of further development.

We estimate the total burden of LC in Colorado to be approximately 9.1%, ranging from 5.6 to 19.1% depending on the probability threshold used. These estimates are similar to estimates provided by recent self-report and records-based studies from similar settings, providing evidence of consistency that bolsters confidence in our methods. For example, a survey-based study comprising 3,000 US adults estimated the LC prevalence to be 7.3% (95% CI 6.1-8.5%) (33), while a study of nearly 450,000 self-reports from US adults estimated the LC prevalence to be 6.4% nationally (95% CI 6.2-6.5%) and in Colorado (95% CI 5.8-7.0%) (21). A study using US medical records and methods similar to ours estimated 10.4% of people with a history of SARS-CoV-2 infection had LC (28), an estimate similar to a Scottish case-control study (34).

Whereas many studies to date have focused on estimating the burden of LC, our study provides unique insights on the potential hidden burdens of LC by providing explicit estimates of undiagnosed LC prevalence, which we estimated to be about 8% (4.5 – 18%), or an estimated 7 undiagnosed cases for every 1 diagnosed case. We have identified two other studies that focused on undiagnosed LC, which was estimated to afflict between 10% and 18% of SARS-CoV-2 positive cases, respectively (26, 28). While in line with the findings of the current study, the fact that both studies relied on prior SARS-CoV-2 infection is a notable potential source of Type II error, due to increases in home testing and immunity-induced asymptomatic or mild cases.

Our ML algorithm highlighted respiratory-related symptoms, malaise and fatigue, nervous system signs and symptoms and higher healthcare utilization as being among the top predictors of LC, all of which were common among both the clinically diagnosed LC cases and the undiagnosed predicted group in our sample. These conditions align with known symptoms of LC, which shows our model is coherent with LC definitions (35–38). High rates of healthcare utilization have also been noted as a defining feature of LC both (39) though our recent related work found that diagnosis may trigger a shift in utilization patterns, with pre-diagnosis more likely to be centered in acute care setting, while outpatient care seeking is more prominent post-diagnosis (22).

Given that XGBoost prediction models generate predicted probabilities based on how much an observation resembles what it has learned to recognize as a “typical” case, it follows that the predicted undiagnosed and diagnosed populations would share similar distributions for many highly ranked predictors, as was highlighted by much of Table 3. Nevertheless, predictors where known LC cases diverge from the predicted undiagnosed population can, with caution, be examined to investigate whether model error, as opposed to a true underlying difference, may be more likely to explain a differential diagnosis status. In our analysis, we found that the predicted undiagnosed group was more likely to be young, and less likely to have dual health insurance (both Medicare and Medicaid) than those with diagnosed LC. As dual coverage implies lower income (Medicaid) plus older age (≥65) and/or disability (Medicare), these findings align with related research highlighting younger age as a risk factor for reduced care seeking behavior in adults (40–42), while improved access to healthcare and insurance is associated with positive health outcomes among older adults (42, 43).

Similarly, both corticosteroids and inhaled bronchodilators were highly ranked positive predictors that were more common among the predicted, undiagnosed group in our final validated model (Table 3). Both prescriptions are frequently used among those with asthma, autoimmune disorders and inflammatory respiratory conditions like COPD, all of which can also increase susceptibility to LC, and/or amplify the symptoms that might necessitate such prescriptions (44–47). It is possible our model is picking up on a pre-diagnosis signal of true underlying LC. Alternatively, the model may be misclassifying LC-like conditions as LC. Taken together, although our model presents risk of misclassification by conflating LC-like conditions with underlying LC, the consistency of many known symptoms of LC in both the diagnosed and predicted undiagnosed group increases our confidence that our model is successfully picking up on true LC signals.

The comparison of our all-predictor model to our sensitivity models yielded some noteworthy discoveries. First, despite expectations that an expert-informed model would achieve optimal performance, the model’s precision was only slightly higher than the all-predictors model (91.9 vs 90.3), while recall was substantially lower (48.0 vs. 56.1). One explanation for this is that the exclusion of several predictors assumed to be irrelevant to a LC diagnosis may have eliminated unique information. As illustrated in Figure 2, some of our most influential predictors in the all-predictors model were negatively associated with a LC diagnosis, suggesting that their utility was primarily in ruling out a LC diagnosis. For example, musculoskeletal pain (MUS010) is a commonly used CCSR code for patients with fibromyalgia which can be particularly difficult to distinguish from LC (5). The fact that MUS010 was among our top predictors and that it was also among the set of negative predictors that were used to rule out LC increases confidence that our model is picking up on subtle signals of true LC. Second, the “Top 50” and “Top 25” model performed similar to the expert-informed model, highlighting that despite some losses to model recall (48.8 and 43.5, respectively), parsimonious models can still perform reasonably well and may be a practical choice when fewer candidate predictors are available or when computing power is more limited. Third, our “binary” model, which used presence/absence (as opposed to continuous or count) for symptoms, utilization metrics and prescriptions, was our model with the lowest recall (recall = 32.0). This suggests that the severity or intensity of health-related indicators also provide important information that can help our model better distinguish LC cases.

The results of our “temporal” model also have important implications. We found that the best performing temporal model iteration was the one that used the latest available data for training (i.e., Folds 2-4) and the earliest available data for testing (i.e., Fold 1), demonstrating better performance than the all-predictors model for precision (90.4 vs. 90.3), recall (65.6 vs. 56.1), F1 score (76.0 vs. 69.2) and AUC-PR (89.9 vs. 87.4). By contrast, the temporal model that used the earliest data for training (Folds 1-3) had the lowest precision (88.8), recall (46.4), F1 score (61.0) and AUC-PR (83.1) of all the temporal models. Given that the COVID pandemic period was characterized by unprecedented and rapidly evolving societal shifts that altered everything from care-seeking behaviors (48), human contact patterns (e.g., the rise of remote school and work) (49, 50), and environmental exposures (e.g., reduced air pollution, reduced pathogen exposure) (51, 52), it is unsurprising that we observed a change in model performance over time. Our study spanned the early pandemic stage to the post-pandemic “new normal” period in which some pre-pandemic conditions have returned along with post-pandemic societal changes (53, 54) and thus may be particularly susceptible to concept drift and temporal degradation (55, 56). Recommended next steps include further study of the potential degradation of the performance of prediction algorithms trained on pandemic era data. As new data becomes available, additional assessments of whether algorithms trained on data from the post-pandemic “new-normal” period demonstrate slower degradation over time will be informative.

## Limitations

There are several limitations in our analysis in addition to those discussed above. First, although our decision to exclude a history of SARS-CoV-2 infection as either a predictor or as a pseudo biomarker of true LC in this study helped to eliminate type II error due to people home-testing or an initial asymptomatic acute infection prior to LC onset, it also made us more susceptible to type I error (i.e., false alarms), a noted phenomenon that is also on the rise due to increasing vigilance and recognition of other LC-like manifestations (e.g., post-acute sequelae of influenza) (57, 58). The absence of a “gold standard” to definitively confirm or rule out LC means that those ascribed a LC U09.9 code include some unknown proportion of misdiagnosed LC, while those without it include an unknown proportion of undiagnosed LC. While we assumed that most mislabeled cases would comprise undiagnosed LC, the prevalence of misdiagnosed LC remains elusive. A range of other respiratory illnesses, inflammatory conditions and autoimmune diseases can lead to LC-like symptoms and utilization patterns (5, 15, 59). Given that most diagnoses are made only after a process of elimination to rule out other LC-like conditions (4, 11, 12), we are confident that most LC diagnoses represent true LC cases, though we acknowledge that the use of the LC U-code does not confirm the presence of true LC.

Second, our analysis does not include uninsured populations or medical treatments not covered by insurance. We assume that the CO APCD, which includes 70% of insured Coloradans (31), is a reasonable approximation of the broader population, an important exception is uninsured populations, who cannot be evaluated using the CO APCD. Likewise, the CO APCD cannot provide information about health-related encounters and purchases that are not covered by insurance (e.g., over-the-counter medicines, elective or experimental procedures, etc.). Another potential source of model error worth noting is that of concept drift, wherein the model’s predictive performance can degrade over time due to broadscale contextual changes (e.g., variants and vaccines, healthcare reform, database coding and record management changes) that render the patterns learned in training data less relevant over time.

## Conclusions

Medical claims data can be leveraged for use in an optimized machine learning algorithm to identify predictors of LC, estimate the burden of undiagnosed LC, and identify factors that make individuals potentially more or less likely to receive a diagnosis, as demonstrated in this study. Such approaches have the potential to fill gaps in our understanding of emerging diseases and conditions, and open new approaches to surveillance of diseases with complex presentations that lack a simple diagnostic test.

## Materials and Methods

### Data sources

For this analysis, we leveraged the CO APCD, which contains claims-based patient demographic and location information, patient symptoms, comorbidities, prescribed medications, and visit characteristics for 71% of the insured Colorado population, comprising over 3.8 million individuals (31, 32). The CO APCD does not include claims from federal programs such as the VA, Tricare, and Indian Health Services; uninsured and self-pay claims; nor the majority of Employee Retirement Income Security Act of 1974 (ERISA)-based self-insured employers.

#### Model training dataset, 2021-2022

We used an extract of 38,064 adults (age 18+) from the CO APCD obtained from CIVHC on December 7, 2023 and enriched with LC cases for model training. The LC-enriched training data, described previously (22), included all clinically diagnosed LC cases (N=12,688) that could be matched to two randomly selected comparators without an LC diagnosis (N=25,376), matching by sex (female, male), age category (18-35, 36-50, 51-64, 65-79, and 80+), and payer (commercial, dual, Medicaid, Medicare Advantage (MA), Traditional Medicare (TM)). We defined dual health insurance as individuals having both Medicare and Medicaid.

LC was identified using the International Classification of Diseases, 10^th^ revision (ICD-10) code U09.9 (post COVID-19 condition, unspecified) (2, 60). A person was classified as having LC if they had at least one instance of a U09.9 code in their record between October 1, 2021 and August 31, 2022. We defined the index date as the date of first appearance of the U09.9 code. Individuals in the comparator group were randomly assigned a pseudo index date to mirror the distribution of index dates among the diagnosed LC cases. For inclusion in this dataset, we required continuous enrollment of at least one year before and after the index or pseudo index date.

The LC enrichment provided a well-characterized set of LC cases for model training and validation, and was developed to address the challenges of modeling rare outcomes (61), given the scarcity of the U09.9 code (<1%) in the total CO APCD population from previous estimates (22). The 1:2 ratio of diagnosed LC cases to comparators allowed us to develop prediction models with greater class balance, enhanced computational efficiency and a reduced noise-to-signal ratio.

#### Target population and unseen data extract, 2022-2023

The target population comprised all adults (18+) who were continuously enrolled in the CO APCD between January 1, 2022 and December 31, 2023 (N = 2,124,386). To estimate the prevalence of undiagnosed LC, we applied our trained model to an unseen data extract from the target population that excluded those already included in the training data (N=33,110) to ensure an unseen, independent cohort for the undiagnosed, predicted prevalence estimation (N = 2,091,276). Prevalence estimation methods are described in detail in “Estimating the prevalence of LC”.

### Long COVID predictors

In our primary model (referred to as the “all-predictors” model), we included 243 candidate predictors of LC across six different categories (clinical symptoms, comorbid conditions, pharmaceutical prescriptions, healthcare utilization, individual demographics, and neighborhood socioeconomic conditions) based on the 2024 consensus definition (3), our prior examination of healthcare utilization patterns of people with an LC diagnosis in Colorado (22), and qualitative interviews with LC providers (62). A full list of predictors is provided in Supplementary Table S2.

Clinical symptoms were classified using the Clinical Classifications Software Refined (CCSR) version 2023.1 (63). CCSR uses ICD-10 codes to define 532 clinically meaningful conditions divided across the 22 body systems. We removed very rare or very common symptoms, excluding CCSR categories that were present in less than 5% or greater than 95% of the diagnosed LC population from the model training dataset, leaving 155 CCSR categories. Each symptom predictor was included as a count of the total number of claims that used each symptom code across the entire study period.

Comorbid conditions were classified using the Charlson Comorbidity Index (CCI) Charlson/Deyo variant (64), which consists of 17 comorbid medical conditions. We also included an indicator for lung cancer, which we separated from cancer comorbidity, and a count variable tallying the number of comorbidities for each patient across the study period.

Healthcare utilization was defined by summing the total number of hospitalizations, emergency department visits, specialist visits, and outpatient office visits for each patient across the study period.

Pharmaceutical prescriptions were classified by mapping national drug codes (NDCs) to drug categories as defined by Veterans Affairs (VA) classes (Supplementary Methods S2: Mapping pharmaceutical prescriptions). We removed very rare and very common prescriptions by excluding VA classes that were present in less than 5% or greater than 95% of the diagnosed LC sample from the model training dataset, resulting in 51 prescription classes. Prescription medications were included as counts of unique occasions on which a drug in a prescription class was prescribed to an individual during the study period. We also included a count of the total number of prescriptions in any class during the study period.

Neighborhood socioeconomic conditions included area-level indicators of socioeconomic (e.g. median household income) and geographic (e.g., distance from the ZIP code centroid to the nearest urgent care) conditions. These were estimated by joining each patient’s APCD-indicated residential ZIP code with the 2020 Agency for Healthcare Research and Quality (AHRQ) Community-Level Health Database (65).

Individual demographic conditions available for this analysis from the APCD included sex, age category, payer, race, ethnicity, and urbanicity. Missingness for individual demographic conditions was included as their own category using a built-in ML missingness function (66). For all other variables, missing data was indicative of having no claims of that particular variable category and was zero-filled.

### Ethics

This research was reviewed by the Colorado Multiple Institutional Review Board (COMIRB) and determined to be exempt under category 4.

### Model training and evaluation

We used a flexible eXtreme Gradient Boosting (XGBoost) approach to generate a prediction model aimed at classifying individuals by their LC diagnostic status (66). XGBoost is an efficient and optimized machine learning algorithm that sequentially builds a collection of simple decision trees, in which each new tree is trained to target and correct the prediction errors made by the previous ones. It has several advantages over traditional regression methods and other ensemble ML methods, including being robust to large and complex datasets, missing data, complex interactions, multicollinearity and heteroscedasticity (67–69).

We randomly assigned each 1:2 matched set of LC cases and their comparators to either a training or validation dataset to yield a 75:25 training-validation split. To handle instances of missing data, we used the XGBoost sparsity-aware split finding function (race was missing for 14.3% of people, and ethnicity for 13.6%; all other variables had <1% missingness, Supplementary Table S1). We used a grid search technique to explore and define key hyperparameter values, specifically the learning rate, gamma, maximum tree depth, minimum child node weight, and the number of decision trees.

#### Model performance

We had higher confidence in the accuracy of our data labels among those with an LC diagnostic code, relative to those without, as we assumed this group largely comprised individuals who managed to overcome the many recognized obstacles to obtaining an LC diagnosis (4). By contrast, we assumed that those without an LC diagnostic code included an unknown mixture of individuals that truly do not have LC and those with undiagnosed LC. In this context, we targeted the optimization of our high confidence data label when assessing model performance, prioritizing model precision (also called positive predictive value) and the Area Under the Precision-Recall Curve (AUC-PR) over recall and the F1 Score. To assess variability in model performance, we conducted 1,000 bootstrap iterations, each time randomly selecting 75% of our sample’s matched sets (each comprising one case and two matched comparators), with replacement. The median and 95% confidence interval (CI) of model precision, recall and F1 score were calculated to summarize variability in model performance across 1,000 bootstrap iterations of our final model (Supplementary Table S3). We evaluated model performance across a range of high probability thresholds and selected a final classification threshold (≥80%; see Supplementary Table S3) that would maximize our understanding of the highest-risk population, while still identifying an acceptably high proportion of all labeled cases (recall), defined here as correctly identifying approximately half of all labeled cases.

#### Predictor importance

We evaluated and visualized variable importance using Shapley Additive exPlanations (SHAP), which provides an estimate of each variable’s average contribution to model predictions: a higher SHAP indicates a greater contribution (70). All variables included in each model were assigned a final variable importance rank based on their mean SHAP, with a rank of 1 indicating the most important variable. The absolute value of each variable’s mean SHAP was summarized across bootstrap iterations to provide estimates of the variability in predictor importance in the primary (“all-predictors”) model (Supplementary Figure S1). Variable importance ranks were also compared between the final all-predictors model and a set of sensitivity models, described below.

#### Sensitivity analyses

We conducted five sensitivity analyses to assess the stability of our primary “all-predictors” model, described in detail in Supplementary Methods S2, and outlined briefly below. First, we tested an “expert-informed” model, which restricted symptoms to 98 CCSR symptoms based on the NASEM definition of LC, a targeted search of the literature, and a review by an interdisciplinary team of LC clinicians. Second, we developed a “binary” model, which used a binary indicator (any/none) for clinical symptoms, pharmaceutical prescriptions, and healthcare utilization to evaluate whether the all-predictors model was biased toward the sickest patients, and if heavy healthcare usage was essential to achieve high predictive performance. Third and fourth, we evaluated the performance of two parsimonious models to assess the feasibility of running a leaner model on a larger dataset and reduce data curation demands. These models included the top 50 and 25 predictors identified in the all-predictors model. Finally, we evaluated model stability and performance over time using a four-fold cross-validation process in our “temporal” model.

#### Estimating the prevalence of undiagnosed LC

To generate estimates of the undiagnosed prevalence of LC in Colorado, we ran our final trained and validated all-predictors model on an unseen data extract from the target population that included adults (18+) who were continuously enrolled in the CO APCD between 2022-2023, excluding those already in the training dataset to give a final sample of N = 2,091,276. Each individual from the target population extract was assigned a predicted probability of LC based on the multifaceted influences of the predictors featured in the final model. We adjusted the output predicted probabilities using a post-hoc correction method developed by King and Zeng 2001 (71) to account for the high prevalence of clinically diagnosed LC in our training data (33%) relative to our best estimate of the real-world prevalence (0.67%) of clinically diagnosed LC in Colorado (not restricting to those continuously enrolled in the CO APCD) (22).

With *ȳ* representing the proportion of LC U09.9 diagnostic codes in our matched training dataset, and *τ* representing the real-world prevalence of LC U09.9 codes, we adjusted the model’s predicted probabilities using the following formula:

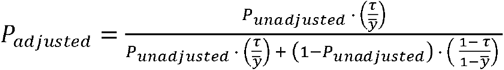

We approximated the final “high-probability” threshold selected during training (80%; see “Model Performance” subsection above, and Supplementary Table S3) using formula (i) to generate a population-relevant adjusted probability threshold to classify individuals as predicted LC, versus not. Those without an LC diagnosis (U09.9 code) who had an adjusted probability at or above the adjusted high probability threshold were assumed to represent the “predicted, undiagnosed” LC population. The final prevalence of undiagnosed, predicted LC was calculated as the number of those at or above the adjusted high probability threshold divided by the total included in the target population extract (N = 2,091,276). This process was repeated for 10-unit increments of unadjusted probability thresholds between 50-90% to estimate the prevalence of undiagnosed LC across the complete range of adjusted high-probability thresholds.

#### Estimating the total prevalence of LC

While our sample is assumed to be robust and generally representative of the broader Colorado population, we note that the observed diagnosed LC prevalence of 1.13% (representing 23,948 cases from our continuously enrolled cohort of 2,124,386 individuals), is higher than a previously estimated diagnosed LC prevalence of 0.67% (representing 26,358 cases across a total of 3,906,402 individuals included in the CO APCD *regardless of the length of enrollment*). To provide a more conservative ratio of the estimated undiagnosed-to-diagnosed LC ratio for our sample, we use the former observed LC prevalence from our continuously enrolled cohort as the basis of our final LC prevalence estimate. We added the observed LC prevalence to the undiagnosed prevalence estimates to generate the total prevalence of LC in CO.

### Comparing diagnosed and undiagnosed LC populations in the APCD

#### Comparison across sociodemographic characteristics

To identify potential inequities in LC diagnosis, we estimated the undiagnosed-to-diagnosed ratio, calculated as the prevalence of predicted, undiagnosed LC, relative to the diagnosed LC prevalence across sociodemographic groups, payer type and access to care. A higher ratio indicates a greater estimated burden of undiagnosed LC (i.e., a larger proportion of cases remain undetected); conversely, a lower ratio indicates a greater portion of the total LC burden has been clinically diagnosed. We used a nonparametric bootstrap procedure with 1,000 resamples of the target population to quantify uncertainty around subgroup-specific ratios. For each bootstrap resample, we calculated diagnosed prevalence among the full resampled target population, and undiagnosed prevalence on the restricted population extract (excluding individuals from the training data). We then calculated the 95% bootstrap confidence interval around the ratio using the 2.5^th^ and 97.5^th^ percentiles of the bootstrap distribution. Those with a diagnosis of LC but missing Sex information comprised <11 individuals and were combined with the Male subgroup to protect data privacy.

## Reproducibility and data availability

Predictive model and prevalence estimation code is available at https://github.com/CRG-CSPH/long_covid. Area-level AHRQ variables were obtained from https://www.ahrq.gov/data/innovations/clh-data.html. The individual-level CO APCD data used for this analysis was provided by CIVHC and under the terms of our data use agreement cannot be distributed. Please contact the corresponding author for details or visit the publicly available CIVHC data portal at https://civhc.org/data/public-data/.

## Author Contributions

ENG: methodology, validation, formal analysis, investigation, writing – original draft, writing – reviewing & editing, project administration; AH: methodology, software, validation, formal analysis, investigation, data curation, writing – review & editing, visualization; EJG: methodology, software, validation, formal analysis, data curation, writing – original draft, writing – review & editing, visualization. RD: resources, data curation, writing – review & editing; RL: resources, data curation, writing – review & editing; SEJ: resources, writing – review & editing, supervision; MCP: conceptualization, methodology, resources, writing – review & editing, supervision, funding acquisition; EJC: conceptualization, methodology, resources, writing – original draft, writing – review & editing, supervision, project administration, funding acquisition.

## Classification

Biological Sciences; Medical Sciences

